# Elevated preoperative suPAR is a strong and independent risk marker for postoperative complications in high-risk patients undergoing major non-cardiac surgery (SPARSE)

**DOI:** 10.1101/2021.05.04.21256448

**Authors:** Athanasios Chalkias, Eleni Laou, Konstantina Kolonia, Dimitrios Ragias, Zacharoula Angelopoulou, Eleni Mitsiouli, Thomas Kallemose, Lars Smith-Hansen, Jesper Eugen-Olsen, Eleni Arnaoutoglou

**Author notes:** **Corresponding author:** Athanasios Chalkias, University Hospital of Larisa, Department of Anesthesiology, C’ Wing, 2^nd^ Floor, PC 41110, Mezourlo, Larisa, Greece.

## Abstract

**Background:** Patients undergoing major surgery are often at risk of developing postoperative complications. We investigated whether the inflammatory biomarker suPAR can aid in identifying patients at high risk for postoperative complications, morbidity, and mortality.

**Methods:** In this prospective observational study (ClinicalTrials.gov identifier: NCT03851965), peripheral venous blood was collected from consecutive adult patients scheduled for major non-cardiac surgery with expected duration ≥2 hours under general anesthesia. Patients fulfilling the following inclusion criteria were included: age ≥18 years and American Society of Anesthesiologists’ physical status I to IV. Plasma suPAR levels were determined using the suPARnostic® quick triage lateral flow assay. The primary endpoint was post-operative complications defined as presence of any complication and/or admission to intensive care unit and/or mortality within the first 90 postoperative days.

**Results:** Preoperative suPAR had an OR of 1.50 (95%CI 1.24-1.82) for every ng/ml increase (AUC 0.82, 95%CI: 0.72-0.91). When including age, sex, ASA score, CRP, and grouped suPAR in multivariate analysis, patients with suPAR between 5.5 and 10 ng/ml had an OR of 12.7 (CI: 3.6-45.5) and patients with suPAR>10 ng/ml had an OR of 20.7 (CI: 4.5-95.4) compared to patients with suPAR≤5.5 ng/ml, respectively. ROC analysis including age, sex, CRP levels, and ASA score and had an AUC of 0.69 (95%CI: 0.58-0.80). When suPAR was added to this Model, the AUC increased to 0.84 (0.74-0.93) (p=0.009).

**Conclusions:** Preoperative suPAR provided strong and independent predictive value on postoperative complications in high-risk patients undergoing major non-cardiac surgery.

## INTRODUCTION

It is estimated that over 300 million operations are performed every year worldwide,^1^ with around 230 million of them being major surgeries.^2,3^ Although the perioperative event rate has declined over the past decades, adverse postoperative complications are a common cause of death and major morbidity in patients undergoing non-cardiac surgery. Complication rates vary among different countries but have been estimated at around 10%,^4^ with 1 in every 13 deaths worldwide occurring within the first 30 days after surgery.^1^ Postoperative complications increase admission to the intensive care unit (ICU) and hospital length of stay, having a significant impact on short- and long-term prognosis and healthcare costs.^2,5^

Risk for postoperative complications depends on many parameters, such as the patient’s preoperative condition, comorbidities, or duration of the surgical procedure, and therefore, the traditional preoperative functional assessment is often insufficient for preoperative risk estimation. Various biomarkers have been suggested with the aim to improve risk stratification beyond that provided by risk scores. However, the most studied biomarkers are limited to predict cardiovascular adverse outcomes only.^6^ A reliable prognostic biomarker that would be able to predict a variety of complications or improve current risk scores and could be incorporated into the process of risk stratification and optimization would be of great value in perioperative planning.

Soluble urokinase plasminogen activator receptor (suPAR) is an immune mediator involved in numerous physiological and pathological pathways, including the plasminogen activating pathway, modulation of cell adhesion, and migration.^7,8^ The specific physiologic role of suPAR is unclear, but its levels in circulation reflect the aggregate activity of the uPAR system with respect to innate immune activity, proteolysis, and extracellular matrix remodeling.^8^ The predictive ability of suPAR has been reported to be equal to or better than other scoring systems or biomarkers in patients presenting to the Emergency Department and is significantly associated with readmission, morbidity, and mortality.^8^ These cumulative data suggest that high suPAR level is a marker of severe disease and may therefore provide benefit for the evaluation of surgical patients to improve risk stratification. The aim of the SPARSE study was to investigate whether the preoperative suPAR level can aid in identifying patients at high risk for postoperative complications, morbidity, and mortality following major non-cardiac surgery.

## METHODS

### Design

This was a prospective observational study conducted in the University Hospital of Larisa from February 2019 to September 2020 and designed in accordance with the declaration of Helsinki.

### Ethics and dissemination

Ethical approval for this study was provided by the Ethical Committee of the University Hospital of Larisa, Larisa, Greece (IRB no. 60580/11-12-2018). The study was registered at ClinicalTrials.gov (NCT03851965) and was performed according to national and international guidelines. Written informed consent was obtained from all patients.

### Patient eligibility

Consecutive patients who were scheduled to undergo elective major non-cardiac surgery with expected duration ≥2 hours under general anesthesia were screened for inclusion. All operative approaches were eligible for inclusion, including open and laparoscopic procedures. Patients fulfilling the following inclusion criteria were included: age ≥18 years and American Society of Anesthesiologists’ (ASA) physical status I to IV.

Exclusion criteria were age <18 years, any infection within the previous four weeks, severe liver disease, patients on renal replacement therapy preoperatively, patients who had previously received a transplant, patients with allergies, inflammatory or immune system disorders, and/or connective tissue disease including rheumatoid arthritis, ankylosing spondylitis, and systemic lupus erythematosus, administration of steroid, antipsychotic, or anti-inflammatory/immunomodulatory medication within the previous 3 months, administration of opioids during the past week, asthma, obesity (BMI ≥30 kg m^-2^), mental disability or severe psychiatric disease, alcohol or other abuse, legal incapacity or limited legal capacity, and subjects currently involved in another study.

### Management of anesthesia and surgical procedures

Endotracheal intubation and anesthetic care were performed according to our institutional routine. Intravenous induction of general anesthesia included i.v. midazolam 0.15-0.35 mg/kg over 20-30 seconds, fentanyl 1 μg/kg, propofol 1.5-2 mg/kg, ketamine 0.2 mg/kg, and rocuronium 0.6 mg/kg. All drugs were prepared in labelled syringes and induction was achieved by administration of a predetermined i.v. bolus dose on the basis of the patient’s weight and/or age. Laryngoscopy and intubation proceeded in a standard fashion, while the position of the endotracheal tube was confirmed by auscultation and capnography/capnometry. The patients were then connected to an automated ventilator (Draeger Perseus A500®; Drägerwerk AG & Co., Lübeck, Germany).

All patients were ventilated using a lung-protective strategy with tidal volume of 7 mL/kg, positive end-expiratory pressure of 6-8 cmH_2_O, and plateau pressures <30 cmH_2_O. Maintenance of general anesthesia included desflurane 1.0 MAC with 40% oxygen and 60% air, while intraoperative fraction of inspired oxygen was adjusted to maintain normoxia. Depth of anesthesia (bispectral index-BIS, Covidien, France) was monitored, with the target ranging between 40 and 60.^9,10^ Normocapnia was maintained by adjusting the respiratory rate as needed, while normothermia (37 °C) was maintained throughout the intraoperative period. All patients were operated by at least one consultant surgeon and a Professor of Surgery.

### Sampling and laboratory measurements

Participants underwent sampling of peripheral venous blood immediately after arrival to the operating room and before induction of anesthesia. Blood samples drawn from all patients were collected in EDTA tubes and were centrifuged at 3.000 x g for 1 min. Plasma suPAR levels were then determined using the suPARnostic® quick triage lateral flow assay (ViroGates, Denmark). According to the manufacturer’s instructions, there is no detectable impact on plasma suPAR concentration when comparing 1 and 10 min of centrifugation.

The suPARnostic® Quick Triage, is an easy-to-use, quantitative test that is based on the lateral flow principle. The device consists of a nitrocellulose membrane with two immobilized antibody zones and a running buffer with gold particles. The quantitative results are read within 20 min by an optical aLF Reader (Qiagen, Germany) with a detection interval of 2-15 ng/mL suPAR.

### Outcomes

The primary endpoint was the presence of complications and/or admission to ICU and/or mortality within the first 90 postoperative days. We used the Clavien-Dindo Classification to assess postoperative complications, morbidity, and mortality in our patients.^11,12^ The Comprehensive Complication Index (CCI®) calculator is an online tool to support the assessment of patients’ overall morbidity (https://www.assessurgery.com/clavien-dindo-classification/). The CCI® is based on the complication grading by Clavien-Dindo Classification and implements every occurred complication after an intervention. The overall morbidity is reflected on a scale from 0 (no complication) to 100 (death). This scoring system offered the advantages of being able to compare results over different time periods within the same institution.^11^ According to related studies, a CCI of 26.2 was set as the cut-off point (equivalent to one grade IIIa complication by the C-D classification), and patients with complications were divided into a high-CCI group (group A, CCI ≥ 26.2) and a low-CCI group (group B, CCI < 26.2) accordingly.^13,14^

### Data collection and monitoring

Data analysis was based on predefined data points on a prospective data collection form. The staff was blinded to measurements until the end of the study and all data were analyzed. Clinical monitoring throughout the study was performed to maximize protocol adherence, while an independent Data and Safety Monitoring research staff was monitored safety, ethical, and scientific aspects of the study. Data collection included demographics, ASA score, anesthesia parameters, general blood count, biochemistry profile, and C-reactive protein (CRP). Considering that the ASA score is not designed to predict mortality, has known inter-rater variation, and offers at least a moderate predictive ability for mortality in multiple surgical settings, we also included ACS-NSQIP and the Charlson Age-Comorbidity Index (Charlson score) in our analysis for the purposes of risk adjustment within research into perioperative outcomes.^15^

### Data management

The goal of the clinical data management plan was to provide high-quality data by adopting standardized procedures to minimize the number of errors and missing data, and consequently, to generate an accurate database for analysis. Remote monitoring was performed to signal early aberrant patterns, issues with consistency, credibility, and other anomalies. Any missing and outlier data values were individually revised and completed or corrected whenever possible.

### Statistical analysis

suPAR was used either as a continuous variable, log2 transformed, or grouped into three groups; ≤5.5 ng/ml; 5.5 - 10 ng/ml; and >10 ng/ml, respectively.^16^ We chose the cut-off of 5.5 ng/ml as this has been previously used in a study of preoperative suPAR levels and post-operative complications^16^ and thus allows for comparison of previous findings. As the chosen cut-off gave a rather large group above 5.5 ng/ml, we chose a second cut-off at 10 ng/ml. There is no specific rationale for this second cut-off, except that suPAR in double digits is often referred to unusually high levels. Association of baseline risk scores (ASA score, ACS-NSQIP, and Charlson Comorbidity Index), CRP(log10), and suPAR with primary endpoint (complications and/or admission to ICU and/or mortality within the first 90 postoperative days) was analyzed with logistic regression models. Both univariable models and multivariable models including age, sex, ASA score, CRP, and suPAR in a single model were fitted. SuPAR was included as continues variable in the univariable analysis and as the 3-level categorical variable in the multivariable analysis. Odds Ratio (OR) with 95% confidence intervals (CI) was reported for the logistic regression models. ROC analysis was carried out using continuous variables to evaluate the predictive level of the baseline risk scores, CRP, and suPAR in relation to the primary endpoint. Analysis was done for each variable separately, as a model with age, sex, ASA score, and CRP and a model with age, sex, ASA score, CRP, and suPAR in order to compare the additional predictive value of suPAR. ROC analysis results are presented as Area Under the Curve (AUC) with CI. The relation between suPAR and CCI was presented graphically. We chose to include 100 individuals as we expected that this number could reveal important associations and generate results to be used for sample size estimation in future largescale studies. The statistical analysis was conducted in SAS Enterprise Guide v. 7.15 HF7 (7.100.5.6177) (64-bit). The main statistics procedure used is proc logistic.

## RESULTS

Of the 100 patients undergoing surgery, 68 (68%) were men and 32 (32%) were women. Median age was 70 years (IQR 62.5-75.5). Our sample included 17 (17%) ASA II, 43 (43%) ASA III, and 40 (40%) ASA IV patients. Baseline characteristics and distribution of baseline parameters according to suPAR predefined levels are shown in Table 1.

**Table 1.** Baseline characteristics and distribution of baseline parameters according to suPAR predefined levels

|  | All | suPAR ≤ 5.5 ng/ml | 5.5 ng/ml < suPAR ≤ 10 ng/ml | 10 ng/ml < suPAR | p value |
| --- | --- | --- | --- | --- | --- |
|  | N=100 | n=27 | n=47 | n=26 |  |
| Age (years), median (q1/q3) | 70.0 (62.5/75.5) | 63.0 (53.0/70.0) | 74.0 (65.0/77.0) | 70.5 (65.0/76.0) | 0.0006 |
| Sex - male | 68 | 16 (23.5 %) | 35 (51.5 %) | 17 (25.0 %) | 0.3803 |
| Sex - female | 32 | 11 (34.4 %) | 12 (37.5 %) | 9 (28.1 %) |  |
| BMI (kg/m<sup>2</sup>), median (q1/q3) | 26.6 (24.7/29.8) | 27.7 (24.9/31.2) | 27.0 (25.1/28.7) | 25.9 (23.3/31.1) | 0.5974 |
| Serum creatinine (mg/dL), median (q1/q3) | 0.80 (0.73/0.96) | 0.75 (0.71/0.90) | 0.80 (0.73/0.91) | 0.94 (0.76/1.21) | 0.0860 |
| CRP (mg/dL), median (q1/q3) | 0.81 (0.24/2.10) | 0.78 (0.24/2.40) | 0.50 (0.14/2.05) | 1.25(0.37/3.16) | 0.1454 |
| Charlson score* (points), median (q1/q3) | 4 (3/6) | 3 (2/4) | 5 (4/6) | 5 (3/6) | 0.0022 |
| ASA classification, median (q1/q3) | 3 (3/4) | 3 (2/4) | 3 (3/4) | 3 (3/4) | 0.0548 |
| ACS-NSQIP (%), median (q1/q3) | 11.8 (7.5/19.9) | 9.3 (4.6/16.7) | 10.8 (8.1/19.7) | 17.3 (10.3/20.6) | 0.0522 |

\*Charlson Age-Comorbidity Index
p-values for sex is Pearson Chi<sup>2</sup>, calculated in SAS-Proc Freq.
All other p-values is Kruskal-Wallis Chi<sup>2</sup>, calculated in SAS-Proc npar1way.

### Logistic regression of baseline risk scores and association with postoperative complications

Postoperative complications are depicted in Table 2. A univariate logistic analysis revealed that ASA score, ACS-NSQIP score, and Charlson Comorbidity Index were associated with postoperative complications with an OR for every increase in score of 1.61 (CI: 0.89-2.90), 0.99 (CI: 0.94-1.04), and 1.34 (CI: 1.04-1.72), respectively.

**Table 2.** Complications according to the type of surgery

| **Type of surgery** | **N** | **Age**<br>**(Median, IQR, min, max)** | **suPAR ng/ml**<br>**(Median, IQR, min, max)** | **CCI-score**<br>**(Median, IQR, min, max)** | **Complications (%)** |
| --- | --- | --- | --- | --- | --- |
| Gastrointestinal | 41 | 68<br>(12/27/89) | 7.9<br>(6.6/1.9/15.1) | 8.7<br>(22.6/0.0/71.7) | 56.1 |
| Gynecological | 5 | 73<br>(15/42/81) | 7.6<br>(3.10/4.20/10.60) | 20.9<br>(20.9/0.0/41.8) | 80 |
| Urological | 16 | 65<br>(12.5/22.0/79.0) | 7.3<br>(5.2/1.9/13.2) | 21.8<br>(18.5/0.00/95.3) | 87.5 |
| Vascular | 32 | 71.5<br>(10.5/24.0/83.0) | 8.3<br>(4.5/1.9/14.3) | 20.9<br>(29.6/0.0/66.9) | 71.9 |
| Other | 6 | 68.5<br>(10.5/24.0/83.0) | 6.5<br>(3.1/2.4/8.2) | 14.8<br>(20.9/0.0/42.4) | 66.7 |

Other types include endocrinological (n=1), thoracic (n=3), gastrointestinal and gynecological (n=1), and thyroidectomy (n=1).

### Association of baseline CRP and suPAR with postoperative complications

We assessed if the baseline levels of CRP and suPAR are associated with the development of postoperative complications. With regard to CRP (entered as a log 10 transformed variable), the OR was 1.33 (CI: 0.71-2.48). Thus, CRP levels were not significantly associated with postoperative complications. In contrast, preoperative suPAR entered as a continuous variable had an OR of 1.50 (CI: 1.24-1.82), thus, for every ng/ml increase in baseline suPAR, the patient had 50% increased odds of developing postoperative complications. To further explore the relationship between continuous suPAR and CCI, these were plotted as shown in Supplementary Figure 1. The AUC of CRP was 0.55 (CI: 0.42-0.68). In contrast, suPAR had an AUC of 0.82 (CI: 0.72-0.91) (Figure 1).

**Figure 1.**
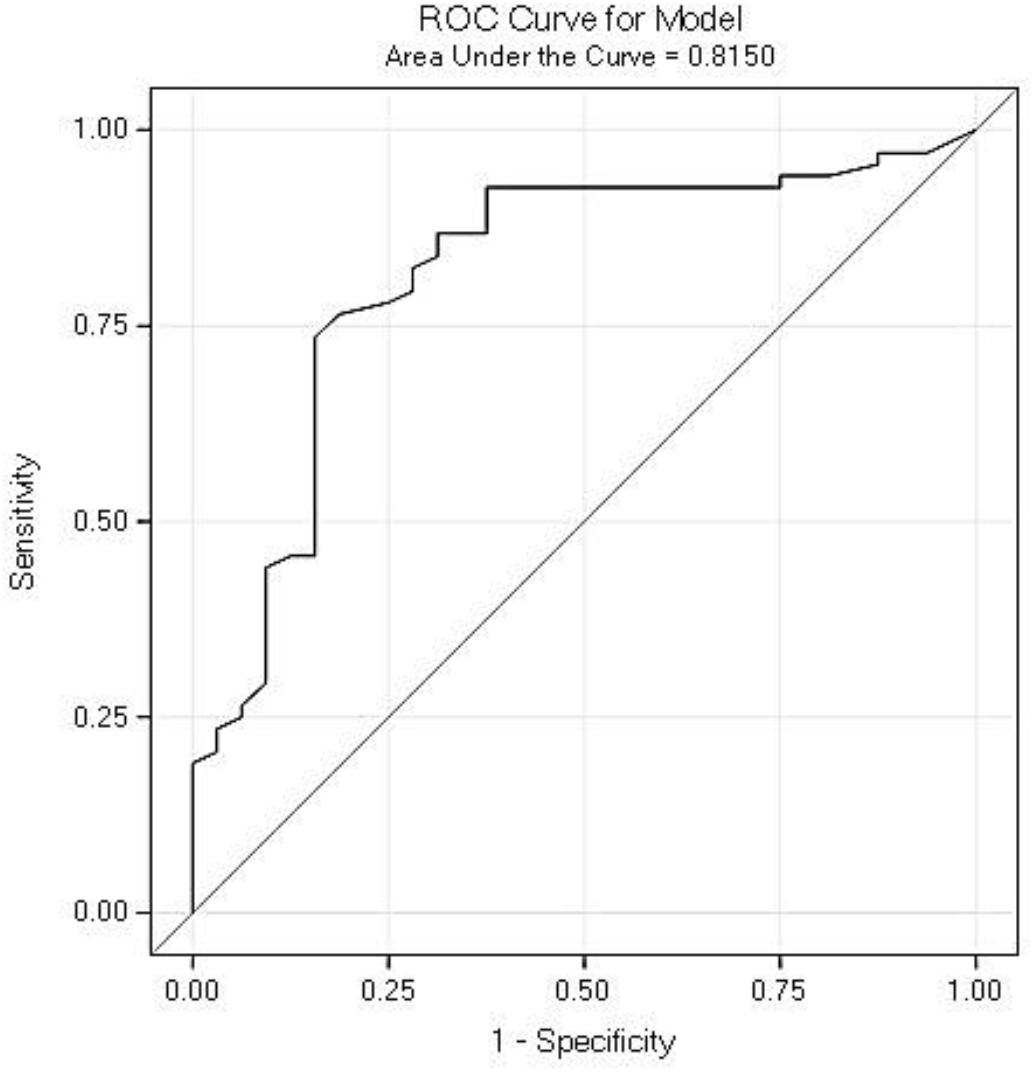
ROC curve of continuous suPAR for complications at day 90. Complications were defined as complications, mortality, or admission to ICU within 90 days after surgery.

### suPAR is strongly and independently associated with postoperative complications

In the multivariate logistic regression analysis, suPAR was included as a 3-level variable (as shown in Table 1): patients with suPAR ≤5.5 ng/ml (n=27), patients with suPAR between 5.5 and 10 ng/ml (n=47), and patients with suPAR >10 ng/ml (n=26). When including age, sex, ASA score, CRP, and grouped suPAR in multivariate analysis, patients with suPAR level between 5.5 and 10 ng/ml had an OR of 12.7 (CI: 3.6-45.5) and patients with suPAR >10 ng/ml had an OR of 20.7 (CI: 4.5-95.4) when compared to patients with suPAR ≤5.5 ng/ml, respectively.

### Combined ROC analysis for prediction of postoperative complications

ROC analysis including age, sex, CRP levels, and ASA score had an AUC of 0.69 (CI: 0.58-0.80), and an AUC of 0.84 (CI: 0.74-0.93) when suPAR was additionally included (Supplementary Table 1), which significantly improved the prediction (p=0.009). The ROC curves are shown in Figure 2.

**Figure 2.**
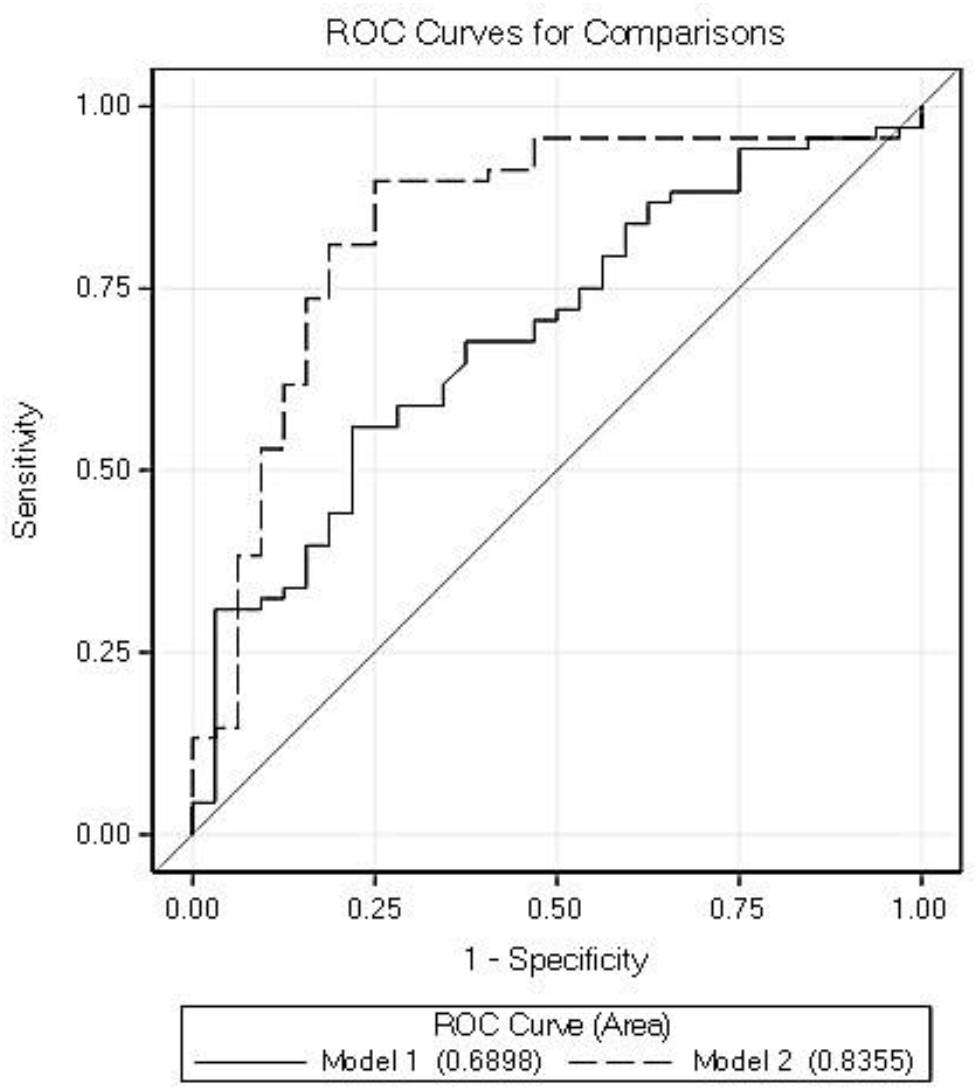
ROC curve for Model 1 (Age, sex, CRP, and ASA score) and Model 2 (Age, sex, CRP, ASA score, and suPAR). Model 2 was significantly better than Model 1.

## DISCUSSION

In this prospective observational study, we report that preoperative suPAR provides strong and independent association with postoperative complications in high-risk patients undergoing major non-cardiac surgery. For every ng/ml increase, a patient has 50% increased odds of developing postoperative complications. Furthermore, the addition of suPAR to a model including age, sex, CRP levels, and ASA score significantly improved the prediction of postoperative complications.

Appropriate risk assessment plays an integral role in optimizing perioperative management to achieve best possible outcomes. However, failure to identify patients at a high risk remains a significant concern and is associated with low-quality postoperative care and increased morbidity and mortality.^17,18^ Risk scores and risk prediction models are used to make a preoperative assessment, but they utilize various predictors and intend to predict different outcomes. Similarly, preoperative biomarkers may improve risk stratification, but their use is currently limited to predicting cardiac complications (e.g., high sensitivity Troponin T or natriuretic peptides).^15^ On the other hand, preoperative resources, such as scheduled ICU admission, would be more appropriately allocated if patient risk could be better assessed. Previous studies have reported that patients undergoing major non-cardiac surgery are at high risk of complications and death,^19-21^ Nevertheless, the development of a universal, rapidly administered, risk assessment tool specific to major surgeries has so far been elusive.

Systemic inflammation plays a major role in the development of cardiovascular and other diseases and its mediators directly injure the cells and/or modulate their response to damage.^22,23^ Consequently, the preoperative inflammatory status can be a major contributor to postoperative organ injury and therefore it is very important for risk stratification.^22,24^ In addition, preoperative neuroendocrine responses to stress can modify the immune function and are associated with adverse outcomes,^25-28^ while several drugs may induce inflammation or exert significant anti-inflammatory properties.^29-31^ Therefore, preoperative suPAR levels may also assist in measuring inflammation, identifying a proinflammatory, inflammatory, or an immuno-suppressive status, which can influence short- and long-term outcome and/or disease progression. Of note, smoking cessation has been reported to lead to a significant (around 1 ng/ml) drop in suPAR within 4 weeks, which in conjunction with the results presented in the current study, may in part explain the importance of smoking cessation before surgery.^32,33^ Also, several proinflammatory conditions are associated with high suPAR levels, such as cardiovascular, autoimmune, and lung diseases, atherosclerosis, cancer, or heart failure, and a high suPAR level has been associated with poor prognosis and complications after surgery in other cohorts.^8,16,34,35^

Several studies have assessed CRP as a preoperative marker, reporting that it may contribute to postoperative complications.^36,37^ However, CRP usually rises in acute inflammatory conditions and due to its short half-life, it is mainly used as a serum marker of acute inflammatory states.^37^ In our study, CRP levels were not associated with postoperative complications, which supports the findings of other authors and the NICE guidelines that do not recommend its use as a preoperative marker.^38,39^ In contrast, we found that for every ng/ml increase in preoperative suPAR, the patient has 50% increased odds of developing postoperative complications. The superiority of suPAR as a prognostic biomarker seems to be largely based on its cellular and molecular effects, with its levels in circulation reflecting the aggregate activity of the uPAR system with respect to innate immune activity and cellular injury.^8,40-42^ Of note, suPAR is a more stable molecule, both *in vivo* and *in vitro*, which makes it a more reliable biomarker for reflecting the overall health condition and state of chronic immune activation of the patient.^43^ On the contrary, CRP is synthesized by the liver only in response to mediators released by macrophages and other cells in acute conditions. Importantly, we found that when including age, sex, ASA score, CRP, and grouped suPAR in multivariate analysis, patients with suPAR level between 5.5 and 10 ng/ml had an OR of 12.7, which increased to 20.7 in patients with suPAR >10 ng/ml.

Today, the ASA classification remains the main tool for risk stratifying surgical patients.^15,16^ However, it has been criticized for its subjectivity, inter-observer variability, and inconsistency.^44^ Also, the ASA classification is related to the anesthesiologist’s knowledge and may be less accurate.^45^ In a previous study, suPAR was significantly associated with the occurrence of postoperative complications and was equally as good as the ASA classification in predicting endpoints. Also, the hazard ratio for 90-day postoperative mortality was 2.5 (95% CI: 1.6-4.0) for every doubling of suPAR level after adjusting for age, sex, and ASA classification.^16^ When these authors combined ASA score and suPAR level, they reported an improved prediction of mortality or complications within 90 days after surgery.^16^ Our study shows a significantly increased prediction of postoperative complications and further strengthen the potential of suPAR as a preoperative risk marker. Based on our findings, and the findings of others,^16,45,46^ suPAR may improve the accuracy of ASA score and could also be used as a central parameter in enhanced recovery after surgery protocols. Of note, the other preoperative risk assessment models used in our study have significant limitations.^15^ The ACS-NSQIP surgical risk calculator can be used for predicting major adverse cardiovascular events, but external validations have been inconsistent and tend to favor a conclusion of inadequate performance.^16,47^ The Charlson scores was not developed to evaluate risk in surgical patients, although the later has been used for this purpose.^15^ These scores also require consideration of numerous variables and are complicated and time-consuming.^16,47^

The study has several strengths. It is a well-designed, prospective observational study that relied on collection of clinical, laboratory, and outcome data, capturing a diverse surgical population. Data collection was systematic and all consecutive patients were enrolled. Another strength is the inclusion of many patients with ASA classifications III and IV, which enhances the discriminatory capabilities of suPAR. Moreover, we assessed suPAR as a part of risk models, which included other risk scores. The main limitation is that it is a single-center study and should be reproduced in a multicenter study to improve general applicability. The number of patients may be relatively small for the different types of surgical interventions, but we revealed important associations and results that can be used in future studies. Future studies should be larger to include more variables in multivariate models. Considering that suPAR levels remain uninfluenced of the surgical trauma,^48^ the use of suPAR in the preoperative stratification process, especially of high-risk patients, represents a promising novel approach.^49^

## CONCLUSION

Preoperative suPAR has a strong and independent association with postoperative complications in high-risk patients undergoing major non-cardiac surgery. For every ng/ml increase, a patient has 50% increased odds of developing postoperative complications. The addition of suPAR to other parameters significantly improved the prediction of postoperative complications.

## Data Availability

Data from SPARSE can be made available upon request through a collaborative process. Please contact the corresponding author for additional information.

## Acknowledgements

The authors would like to thank the medical and nursing stuff of the Department of Anesthesiology, University Hospital of Larisa, for their assistance during the study period.

## Funding

No funding received.

## Conflicts of interest

Jesper Eugen-Olsen is a co-founder, shareholder and CSO of ViroGates A/S and is mentioned inventor on patents on suPAR owned by Copenhagen University Hospital Hvidovre, Denmark. All other authors report no conflicts of interest.

## Author Contributions

Dr. Athanasios Chalkias designed the study. Dr. Athanasios Chalkias, Dr. Eleni Laou, Dr. Konstantina Kolonia, Dr. Dimitrios Ragias, Dr. Zacharoula Angelopoulou, Mrs. Eleni Mitsiouli, and Dr. Eleni Arnaoutoglou collected the data and performed quality control. Dr. Athanasios Chalkias, Dr. Thomas Kallemose, Dr. Lars Smith-Hansen, and Dr. Jesper Eugen-Olsen analyzed the data. Dr. Athanasios Chalkias and Dr. Jesper Eugen-Olsen provided expert interpretation of the findings. Dr. Athanasios Chalkias wrote the first draft of the manuscript. All coauthors provided critical revisions to the manuscript. All coauthors had full access to the data and take responsibility for the integrity and accuracy of the data analysis. All authors approved the final version of the manuscript.

## Notes

### Clinical Trial

NCT03851965

