## Supplementary File for "Elevated preoperative suPAR is a strong and independent risk marker for postoperative complications in high-risk patients undergoing major non-cardiac surgery (SPARSE)"

**Supplementary Figure 1.** Correlation plot showing suPAR plottet against complication score. suPAR and complications were entered as continuous variables.

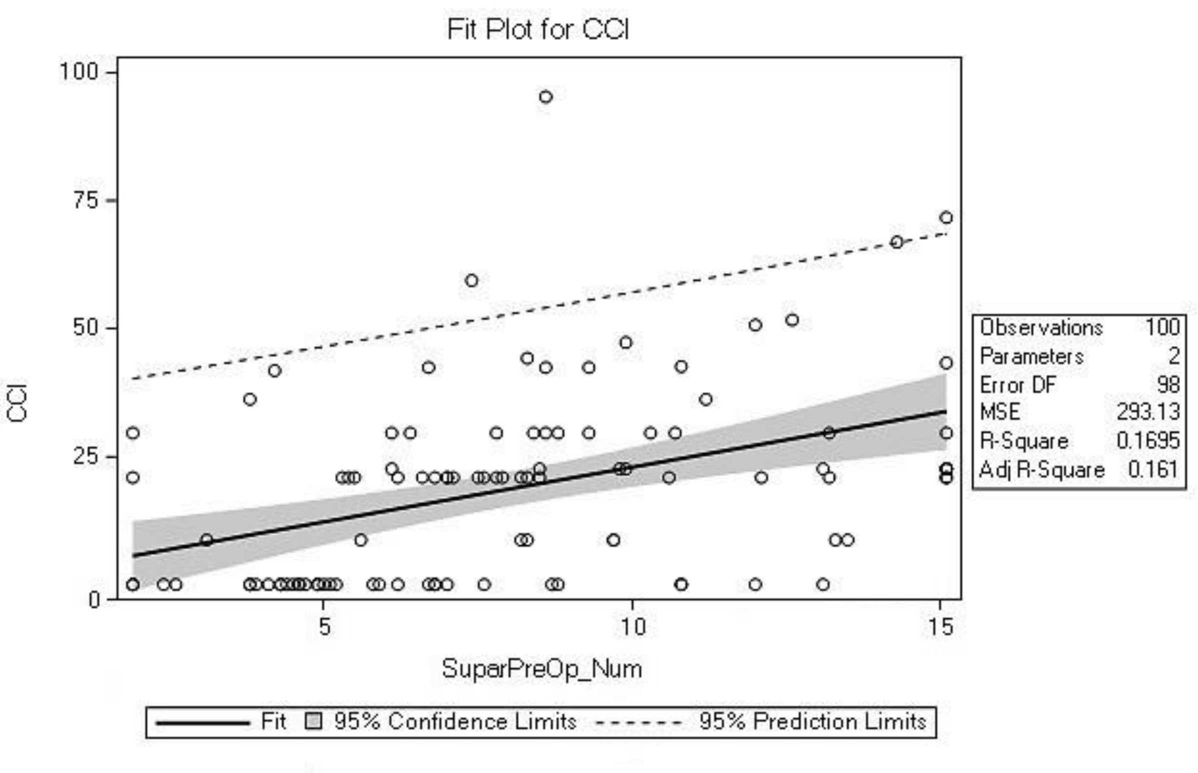

**Supplementary Table 1.** ROC Association Statistics for Model 1 and Model 2

| | | *Mann-Whitney* | | | | | |
|---|---|---|---|---|---|---|---|
| | | *Standard* | *95% Wald* | | | | |
| *ROC Model* | *Area* | *Error* | *Confidence Limits* | | *Somers' D* | *Gamma* | *Tau-a* |
| *Model 1* | 0.6898 | 0.0557 | 0.5807 | 0.7989 | 0.3796 | 0.3799 | 0.1669 |
| *Model 2* | 0.8355 | 0.0475 | 0.7423 | 0.9286 | 0.6710 | 0.6710 | 0.2949 |
